# Altered blood microbiome in patients with HCV-related decompensated cirrhosis

**DOI:** 10.1101/2024.05.06.24306779

**Authors:** Oscar Brochado-Kith, Marta Rava, Juan Berenguer, Juan González-García, David Rojo, Cristina Díez, Victor HontaÑon, Ana Virseda-Berdices, Luis Ibañez-Samaniego, Elba Llop-Herrera, Antonio Olveira, Leire Perez-Latorre, Coral Barbas, Amanda Fernández-Rodríguez, Salvador Resino, María Angeles Jiménez-Sousa, ESCORIAL Study Group

## Abstract

**Background:** Altered bacterial translocation is associated with transitioning from compensated to decompensated cirrhosis. Thus, we aimed to study differences in the blood microbiome of HCV-infected patients with and without hepatic decompensation.

**Methods:** We conducted a cross-sectional study in patients with advanced HCV-related cirrhosis with or without human immunodeficiency virus (HIV) infection (n=88). MiSeq Illumina technology for bacterial 16S rRNA sequencing was used. Non-targeted metabolomics was performed by GC-MS and LC-MS ESI+ and ESI-.

**Results:** Patients with decompensated cirrhosis had lower levels of richness (Chao1), and alpha diversity (Shannon and Simpson indexes) at phylum level, than patients without decompensation. Likewise, we observed significant differences in beta diversity between groups at phylum, class and order levels, being lower in decompensated cirrhotic patients. Higher relative abundance of Proteobacteria (Fold Change (FC)=1.54, p=0.012), Alphaproteobacteria (FC=1.57, p=0.016) and Sphingomonadales (FC=1.61, p=0.050) were significantly associated with hepatic decompensation. The phylum Proteobacteria was positively correlated with ethanolamine and oleic acid (p=0.005 and p=0.004, respectively) and negatively with p-cresol (p=0.006). In addition, the order Sphingomonadales was also negatively correlated with p-cresol (p=0.001).

**Conclusions:** Blood microbial diversity was significantly decreased in patients with decompensated cirrhosis, who presented an enrichment of Proteobacteria, Alphaproteobacteria, and Sphingomonadales, compared to patients with compensated cirrhosis.

## Background

Hepatitis C virus (HCV) infection is considered one of the most notable causes of chronic liver disease worldwide. Without antiviral therapy, about 5-20% of HCV-infected patients will develop cirrhosis, increasing the risk of liver failure, hepatic decompensation, hepatocellular carcinoma (HCC), and death. Hepatic decompensation is considered the most important cause of hospitalization among cirrhotic patients and a major risk factor for death [1]. Decompensated cirrhosis has an annual mortality rate of 57%, and 30% mortality is associated with acute decompensating events [2]. Traditionally, non-invasive indexes have been used to triage patients with advanced liver diseases. Among them, the Child-Turcotte-Pugh (CTP) score is an essential indicator of severity, determining the cirrhosis status and predicting morbidity and mortality.

The gut microbiota and bacterial translocation may influence liver disease progression in patients with viral hepatitis [3]. Likewise, gut microbiota dysbiosis and sustained bacterial translocation have been observed among people living with human immunodeficiency virus (HIV) despite the use of antiretroviral therapy (ART) [4]. In this setting, bacterial overgrowth increases intestinal permeability, and defects in gut-associated lymphatic tissue in patients with advanced liver disease promote impaired bacterial translocation, which leads to severe liver damage by several mechanisms related to persistent immune activation and inflammation [5]. Thus, altered bacterial translocation is associated with transitioning from compensated to decompensated cirrhosis [6].

In this context, the presence in the blood of certain bacteria or bacterial products, and the finding of specific microbiome patterns, could constitute an innovative approach for a better understanding of the pathogenesis of advanced HCV-related cirrhosis. So far, scarce studies have investigated the blood microbiome in decompensated cirrhotic patients. Trakova *et al.* [7] found a markedly higher number of bacterial species than control individuals. Alvarez-Silva *et al.* [8] described a relationship between the microbiota composition and systemic inflammation in the blood. However, to our knowledge, none of the previous studies focused on studying the role of the blood microbiome among HCV-infected patients with hepatic decompensation.

### Objective

We aimed to study differences in the blood microbiome of HCV-infected patients with and without hepatic decompensation.

## Methods

### Study subjects

We performed a cross-sectional study in patients with advanced HCV-related cirrhosis with or without HIV, recruited at four tertiary referral hospitals in Madrid (Spain) between January 2015 and June 2016. The study received the approval of the Research Ethics Committee of the Instituto de Salud Carlos III (CEI42_2020, CEI41_2014) and was carried out following the Declaration of Helsinki. All participants of the study gave their written informed consent.

The selection criteria were: 1) demonstrable active HCV infection by polymerase chain reaction (PCR); 2) advanced cirrhosis defined by any of the following criteria: i) prior history of ascites, bleeding esophageal varices, or hepatic encephalopathy; ii) liver stiffness ≥25 kPa; iii) CTP ≥7); and/or iii) clinically significant portal hypertension defined as an hepatic venous pressure gradient (HVPG) ≥ 10 mmHg; 3) available CTP score; 4) available blood sample for microbiome analysis. HIV/HCV-coinfected patients had a stable ART for over six months and undetectable plasma HIV viral load (<50 copies/mL).

### Samples

Approximately 30-40 mL of whole blood was collected from each patient in EDTA tubes, which were sent to the HIV Biobank (http://hivhgmbiobank.com/?lang=en), where they were processed and stored at -80 °C until use. The epidemiological and clinical variables were collected using an online form within each center, which fulfilled data confidentiality requirements.

### Outcome variable

The outcome was hepatic decompensation, defined as a CTP score ≥ 7. CTP score was calculated from five routine laboratory parameters and clinical measures of liver disease (serum albumin, total bilirubin, international normalized ratio (INR), ascites, and hepatic encephalopathy) (https://www.hepatitisc.uw.edu/page/clinical-calculators/ctp).

### Blood microbiome

Whole blood samples were used for DNA extraction and subsequent 16S targeted metagenomic sequencing in a strictly controlled environment. It was carried out at Vaiomer (Toulouse, France), a biotech company expert in tissue and blood microbiota, which uses a rigorous contamination-aware approach, described and discussed elsewhere [9–11].

DNA was extracted from 50 µl of each whole blood sample using an optimized tissue-specific technique, as previously described [9, 11]. The quality and quantity of extracted nucleic acids were controlled by gel electrophoresis and NanoDrop 2000 UV spectrophotometer (Thermo Scientific).

Library preparation was performed by two-step PCR amplification using 16S universal primers targeting the V3–V4 region of the bacterial 16S ribosomal DNA (rDNA), as described previously [10]. The resulting amplicon of approximately 467 base pairs was sequenced using 2 x 300 paired-end MiSeq kit V3. For each sample, a sequencing library was generated by the addition of sequencing adapters. The detection of the sequencing fragments was performed using MiSeq Illumina technology.

The targeted metagenomic sequences from microbiota were analyzed using the bioinformatics pipeline established by Vaiomer based on the Find, Rapidly, OTUs with Galaxy Solution guidelines. Briefly, after demultiplexing the barcoded Illumina paired reads, single-read sequences were cleaned and paired for each sample independently into longer fragments. Operational taxonomic units (OTUs) were produced via single-linkage clustering using the Swarm algorithm and its adaptive sequence agglomeration [12]. The taxonomic assignment was performed against the Silva v132 database to determine taxonomic profiles. The following specific filters were applied for this analysis to obtain the best results: (1) the last ten bases of reads R1 were removed; (2) the last 40 bases of reads R2 were removed; (3) amplicons with a length of <350 or >500 nucleotides were removed; (4) OTUs with abundance <0.005% of the whole dataset abundance were removed. To ensure the low impact of the potential DNA contamination from the environment and especially from reagents, negative controls were added and carried over throughout the 16S rRNA gene sequencing pipeline. Negative controls were performed for the DNA extraction and amplification steps with molecular grade water as starting material. Positive controls with a mock community were also added during the sequencing library preparation.

### Non-targeted metabolomics

Firstly, methanol was mixed with plasma samples (3:1, v/v) for viruses’ inactivation. Then, samples were vortexed (15 sec), maintained cold for 5 min, centrifuged (16000 g, 20 min, 4°C), and frozen (−80°C) before being sent to the Center for Metabolomics and Bioanalysis (CEU-San Pablo University, Pozuelo de Alarcón, Spain). On the day of analysis, the samples were correctly processed and subsequently analyzed by gas chromatography-mass spectrometry (GC-MS) and liquid chromatography-mass spectrometry (LC-MS) with positive and negative electrospray ionization (ESI). Quality controls were prepared by pooling and mixing equal volumes of each corresponding sample independently for each analytical platform (full description in **Supplementary_file_1**).

For the deconvolution and identification process in GC-MS, MassHunter Quantitative Unknown analysis (B.07.00, Agilent) was used. Alignment was carried out with MassProfiler Professional software (version 13.0, Agilent), and Masshunter Quantitative Analysis (version B.07.00, Agilent) was used for peak integration. In LC-MS, the Molecular Feature Extraction and the Recursive Feature Extraction algorithms in the MassHunter Profinder software (B.08.00, Agilent) were used for deconvolution, peak integration, and alignment of the raw data (more details are available in **Supplementary_file_1**).

### Statistical analysis

For the descriptive study, quantitative variables were expressed as median (interquartile range) and categorical variables as absolute count (percentage). When comparing data between groups, we used Fisher’s exact test for categorical and unpaired Wilcoxon rank-sum test for continuous variables.

Regarding the metagenomics data, richness (Chao1 estimator), alpha diversity (Shannon and Simpson index), and beta diversity (Weighted Unifrac distance, Bray-Curtis dissimilarity, Jaccard index) of bacterial communities were calculated (Vegan package V. 2.5-7). Richness and alpha diversity indexes were analyzed in association with hepatic decompensation, here considered as an independent variable, by the Wilcoxon rank-sum test and multivariable generalized linear model (GLM) with a gamma distribution, unadjusted and adjusted by HIV coinfection (included as covariate in the model). Besides, we compared beta diversity between groups by using principal coordinates analysis (PCoA) plots and permutational MANOVA.

For analyzing differences in the relative abundance of OTUs between groups, we first normalized the initial data at each taxonomic level and filtered those taxa that did not reach the minimum abundant level for the analysis (at least appeared in 10% of the samples). Subsequently, we carried out univariable and multivariable analyses. Univariable analysis was performed using the Wilcoxon rank-sum test, a non-parametric test that compares the relative abundances of bacterial taxa between groups. The false discovery rate (FDR) was applied to account for multiple comparison, and adjusted p-values (q-values) were calculated according to the Benjamini-Hochberg method. Those taxa with p<0.05 and q<0.150 were included in the multivariable models. Multivariable analyses were carried out using the aldex.glm function from the ALDEx2 package (v. 1.22.0), and HIV coinfection was included as a covariate.

Finally, we selected the bacterial taxa significantly different in the relative abundance between groups, and we estimated the correlation with plasma metabolomic data from GC-MS, LC-MS ESI+, and LC-MS ESI-using the Spearman rank order correlation test. The correlations were considered relevant when r>0.3 or r<–0.3 and q<0.150.

All the analyses were carried out by using R statistical package (R Foundation for Statistical Computing, Vienna, Austria, v. 4.0.5).

## Results

### Characteristics of the study population

A total of 88 patients were included, 61 with HIV, whose characteristics stratified by their stage of cirrhosis (compensated vs. decompensated) are shown in **Table 1**. Among patients with compensated cirrhosis (n=75), the mean age was 53.1 years, and 65% were male. Patients with decompensated cirrhosis (n=13) had a mean age of 52.1 years, and 92.3% were male. Of note, decompensated patients were less frequently coinfected with HIV (*p* =0.019).

**Table 1.** Clinical and epidemiological characteristics of patients with HCV-related advanced cirrhosis.

| Characteristic | Compensated | Decompensated | P-value |
| --- | --- | --- | --- |
| **No.** | 75 | 13 |  |
| **Age (years)** | 53.1 [50.5-56.1] | 52.1 [46.6-56.4] | 0.301 |
| **Gender (male %)** | 49/75[65.33%] | 12/13[92.3%] | 0.058 |
| **BMI (kg/m<sup>2</sup>)** | 24.2[22.1-27.7] | 26.7[23.9-27.9] | 0.198 |
| **Smoker** |  |  | 0.389 |
| Never | 16/75[21.3%] | 1/13 [7.7%] |  |
| > 6 months | 17/75 [22.7%] | 5/13 [38.5%] |  |
| Currently | 42/75 [56%] | 7/13 [53.9%] |  |
| **Alcohol intake** |  |  | 0.376 |
| Never | 39/75 [52.0%] | 5/13 [38.5%] |  |
| > 6 months | 31/75 [41.3%] | 6/13 [46.2%] |  |
| Currently | 5/75 [6.7%] | 2/13 [15.4%] |  |
| **IVDU** | 44/75[58.7%] | 8/13[61.5%] | 0.999 |
| **HCV genotype 1** | 51/75[68.0%] | 7/11[57.1%] | 0.743 |
| **HCV viral load (log10)** | 6.2[5.7-6.6] | 5.79 [5.2-6.2] | 0.053 |
| **Previous anti-HCV treatment** | 36/75[48.0%] | 7/13 [53.9%] | 0.770 |
| **Statins** | 9/75[12.0%] | 0/13 [0%] | 0.345 |
| **Coinfection (HIV)** | 56/75[74.7%] | 5/13 [38.5%] | **0.019** |
| **CD4<sup>+</sup>/mm<sup>3</sup>** | 446.5[239.3-719.5] | 378.0[164.0-685.0] | 0.502 |
| **Nadir CD4 cells** | 119.0[63.0-232.5] | 207.0[144.0-300.0] | 0.131 |
| **AIDS** | 37/56[66.1%] | 3/5[60.0%] | 0.999 |

**Statistics:** Continuous variables were expressed as median [interquartil range], and p-values were calculated by the Wilcoxon rank-sum test. Categorical variables were expressed as absolute count [percentage], and p-values were calculated by the Fisher exact test.
**Abbreviations:** CTP, Child-Turcotte-Pugh; BMI, Body mass index; IVDU, Intravenous drugs user; HCV, Hepatitis C virus; HIV, Human immunodeficiency virus; AIDS, Acquired immune deficiency syndrome.

### Microbiome analysis

#### A) Richness and alpha diversity

Decompensated patients showed lower richness at phylum (Chao1, p=0.032) and class levels (Chao1, p=0.020) than compensated patients (**Figure1**). Regarding alpha diversity, decompensated patients showed lower levels of Shannon (p=0.005) and Simpson indexes (p=0.003) at the phylum level than compensated patients (**Figure1**).

**Figure 1.**
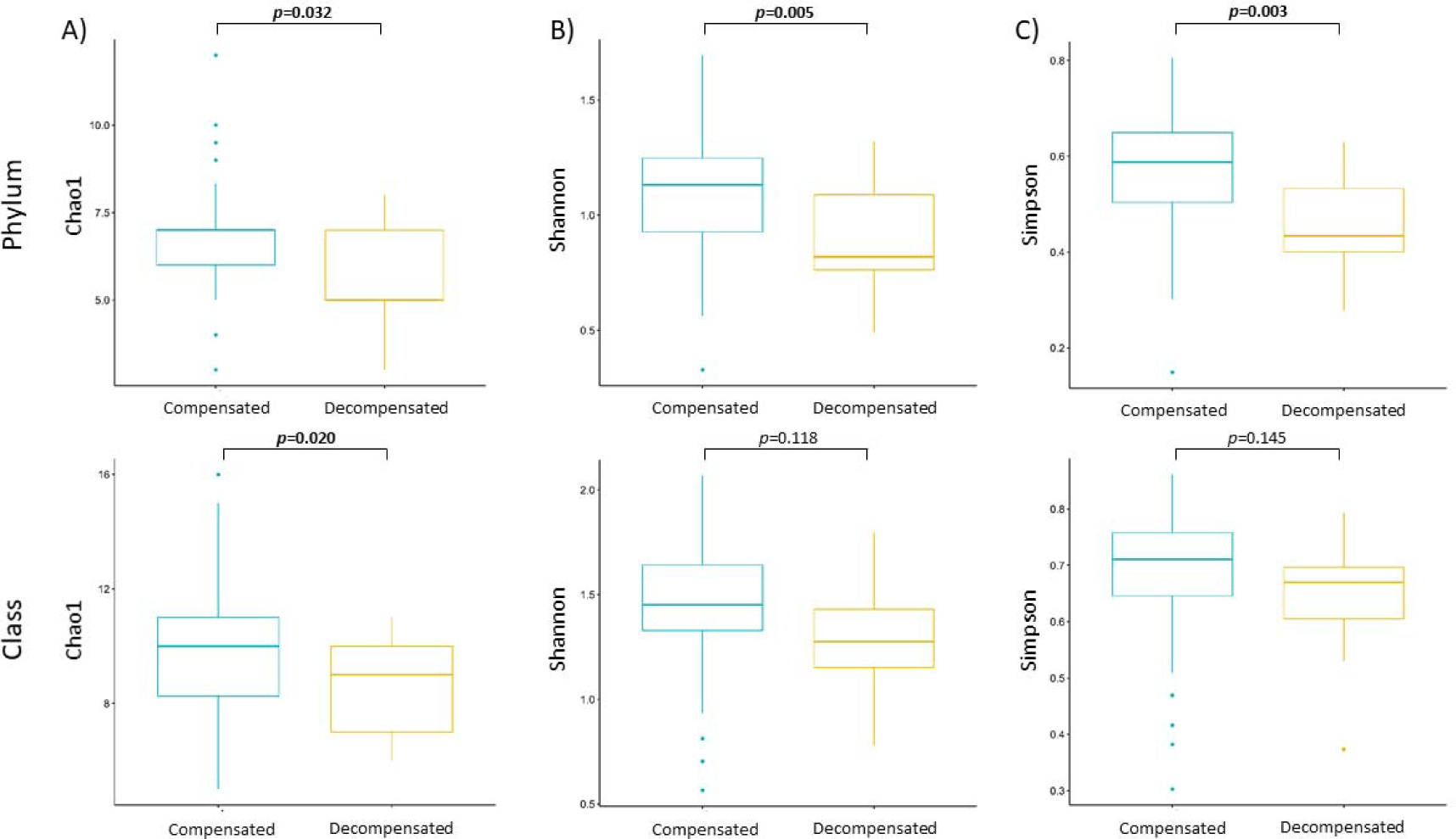
Representation of richness and alpha-diversity indexes for patients with compensated and decompensated cirrhosis. A) Chao1 estimator: it is calculated by taking the number of different species and also considering as relevant the number of species that were detected once (singletons) or twice (doubletons). B) Shannon index: it is based on the formula to describe entropy and how difficult it is to predict the next microbe detection when the diversity is high. C) Simpson index: it measures the probability that two individuals randomly selected from a sample will belong to the same taxa. Differences between groups were calculated by using Wilcoxon signed-rank test and expressed as p-values.

After adjustment for HIV coinfection in a gamma GLM model, we found that decompensated patients had lower levels of Chao1 estimator at the phylum (adjusted arithmetic mean ratio (aAMR)=0.85, p=0.021) and class level (aAMR=0.85, p=0.019) than compensated patients. Regarding alpha diversity, decompensated patients showed lower levels of Shannon (aAMR=0.80, p=0.005) and Simpson indexes (aAMR=0.83, p=0.006) at the phylum level than compensated patients (**Supplementary_file_2)**.

#### B) Beta diversity

Beta-diversity was significantly different between compensated and decompensated patients, finding lower beta-diversity at the phylum level (Jaccard: p=0.049), class level (Weighted Unifrac: p=0.040, Bray-Curtis: p=0.017, Jaccard: p=0.028), and order level (Weighted Unifrac: p=0.016, Bray-Curtis: p=0.047) in decompensated patients, as determined using PCoA plots and permutational MANOVA. In addition, these significant differences were maintained when models were adjusted by HIV coinfection at the phylum level (Jaccard: adjusted p=0.046), class level (Weighted Unifrac: adjusted p=0.033, Bray-Curtis: adjusted p=0.018), and order level (Weighted Unifrac: adjusted p=0.019, Bray-Curtis: adjusted p=0.048) (**Figure2, Supplementary_file_3**).

**Figure 2.**
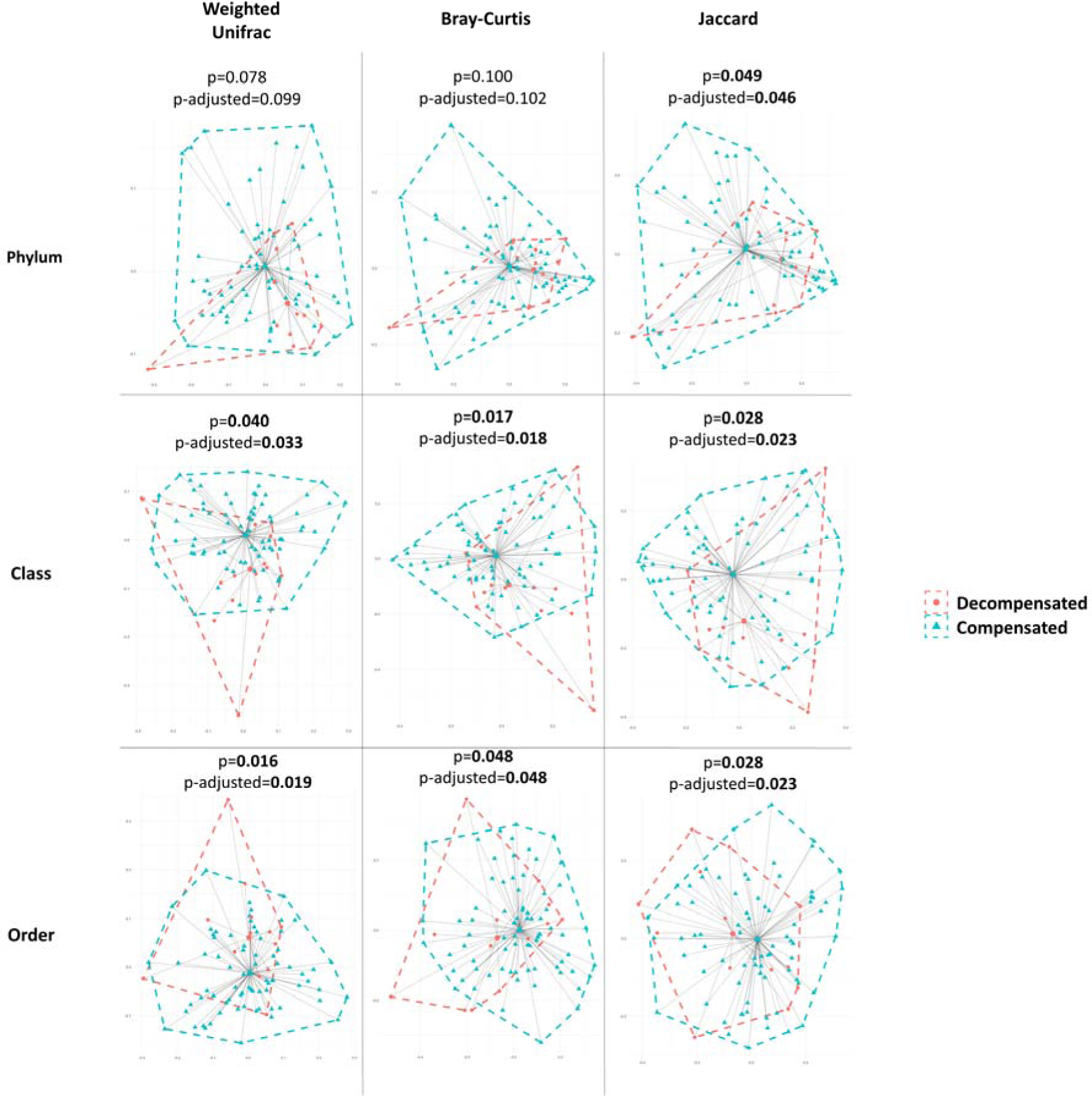
Principal coordinates analysis (PCoA) representing the beta diversity for patients with compensated and decompensated cirrhosis. Statistics: P-values were calculated from a permutational univariate and multivariate analysis of variance adjusted for HIV-coinfection.

#### C) Relative abundances

We assessed the differences in the relative abundance at all taxonomic levels between decompensated and compensated patients. Decompensated patients had a higher relative abundance of the phylum Proteobacteria (p=0.004), the class Alphaproteobacteria (p≤0.001), the orders Sphingomonadales (p≤0.001) and Oceanospirillales (p=0.047), families *Sphingomonadaceae* (p=0.002), *Paenibacillaceae* (p=0.038) and *Microbacteriaceae* (p=0.043), and genus *Bradyrhizobium* (p=0.001), *Sphingomonas* (p=0.009) and *Rhodococcus* (p=0.027) than compensated patients. However, only the phylum Proteobacteria (q=0.037), the class Alphaproteobacteria (q=0.006), the order Sphingomonadales (q=0.050), family *Sphingomonadaceae* (q=0.090) and genus *Bradyrhizobium* (q=0.087) were considered relevant after adjusting for multiple comparisons (FDR, q<0.150) (**Table 2**). When HIV coinfection was taken into account for adjusting the models by ALDEx2, a higher relative abundance of Proteobacteria (p=0.012), Alphaproteobacteria (p=0.016), and Sphingomonadales (p=0.050) remained significantly associated with hepatic decompensation. Boxplots for the relative abundances of these significant bacterial taxa are shown in **Supplementary_file_4**.

**Table 2.**
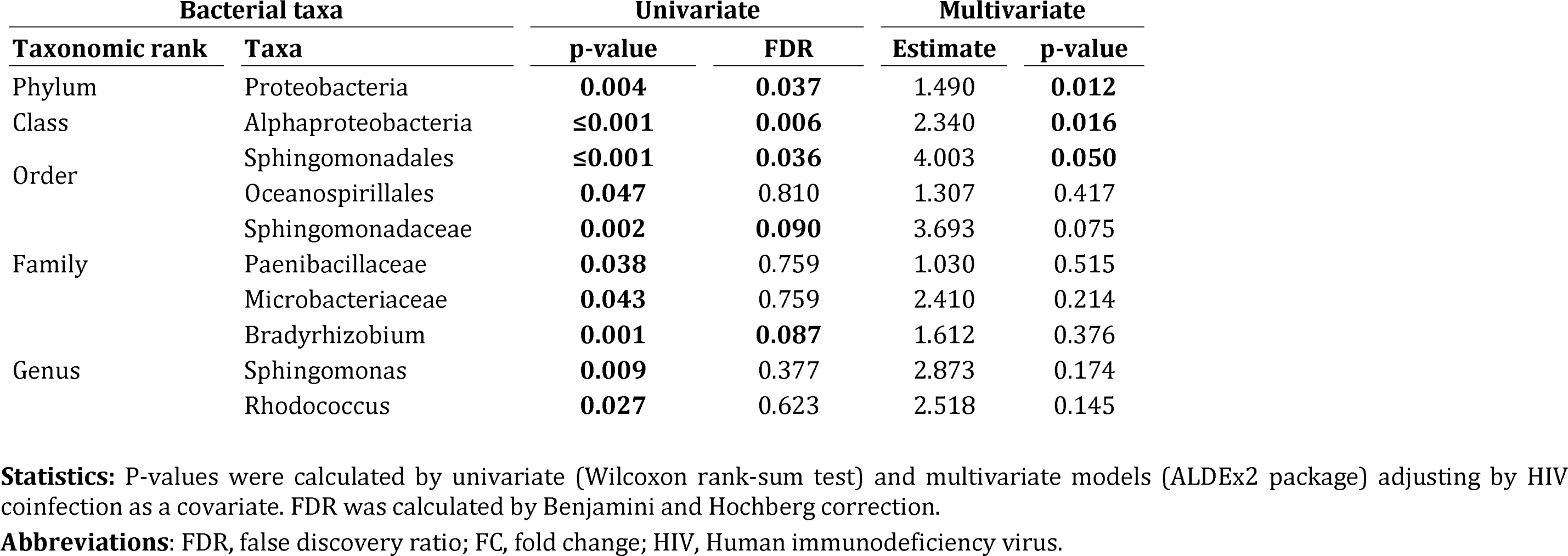
Association between relative abundance of blood bacterial taxa and HCV-related decompensated cirrhosis.

### Correlation analysis with metabolomic data

The correlation between bacterial taxa related to hepatic decompensation and metabolomic data was studied. Regarding GC-MS data, we observed a significant positive correlation of the phylum *Proteobacteria* with ethanolamine and oleic acid (r=0.330, p=0.005, q=0.115 and r=0.302, p=0.004, q=0.115, respectively) and a significant negative correlation with p-cresol (r=-0.308, p=0.006, q=0.115). In addition, the order *Sphingomonadales* was negatively correlated with p-cresol (r=-0.354, p=0.001, q=0.112) (**Figure3, Supplemental File 5**). Regarding LC-MS data, we found a significant negative correlation between the order *Sphingomonadales* and p-cresol (r=-0.367, p=0.001, q=0.122) (**Figure3, Supplementary_file_5**).

**Figure 3.**
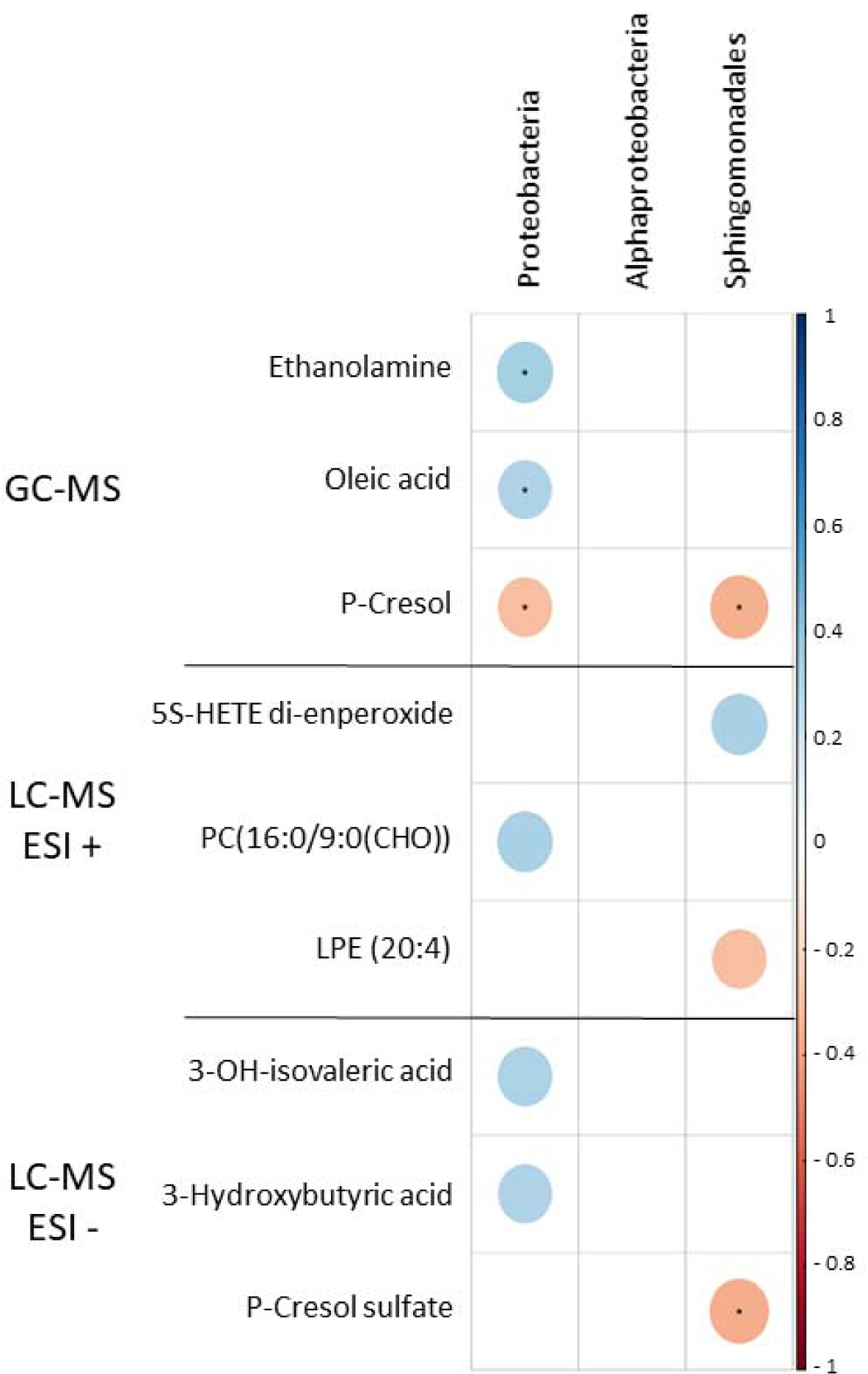
Correlation between blood significant bacterial taxa and plasma metabolites. Statistics: Direct correlations are shown in blue while inverse correlations are shown in red. Asterisks indicate those correlations with a value of p-value<0.05 and q-value≤ 0.15 (q-values (FDR) were calculated from Spearman correlation p-values.

## Discussion

We compared the blood microbiome between HCV-infected patients with decompensated and compensated cirrhosis, finding lower alpha and beta diversity richness in the first ones. We also found that the relative abundance of Proteobacteria, Alphaproteobacteria, and Sphingomonadales was higher in decompensated patients. The dysbiosis of the blood microbiome was accompanied by changes in metabolites related to liver reparation processes and microbiota fermentation.

Although several studies have previously investigated the blood microbiome in cirrhosis, few articles have described its role among HCV-infected cirrhotic patients. Recently, Gedgaudas *et al.* found that circulating microbiome profiles in patients with cirrhosis were distinct from those of healthy individuals, with enrichment of the genera *Bacteroides, Escherichia/Shigella,* and *Prevotella* in patients with portal hypertension [13]. However, this study included patients with cirrhosis of different etiologies and analyzed plasma instead of whole blood [13]. Traykova *et al.* studied the blood microbiome in nine patients with decompensated cirrhosis compared to controls, finding that the bacterial microbiome is related to changes in systemic vascular resistance and cardiac outcomes [7]. However, they only screened for 53 specific bacteria from the gut in the blood [7]. Moreover, our group previously found an association between specific bacterial taxa before HCV therapy, such as *Corynebacteriales* and *Massilia,* and a decrease in HVPG in patients with HCV-related cirrhosis after direct-acting antiviral therapy for HCV [14]. Thus, to our knowledge, this is the first study in which the circulating microbiome from patients with compensated and decompensated HCV-related cirrhosis has been compared, providing new insight into the potential mechanisms underlying this liver injury status.

In our study, patients with decompensated cirrhosis had lower richness and alpha diversity for different taxonomic ranks than patients with compensated cirrhosis. These findings are consistent with previous data supporting that less diverse gut microbiome ecosystems are associated with poorer body health status, probably because reduced species diversity leads to lower efficient systems [15]. Regarding viral hepatitis and gut microbiome, a less diverse microbiome has been previously described in the gut of cirrhotic patients with hepatitis B and C compared to healthy controls, finding significant differences in the microbiota community between groups [16, 17]. Moreover, an improvement in gut microbiota diversity was also observed in responders achieving a sustained virological response after anti-HCV therapy; however, patients with more advanced fibrosis stages showed lower improvement [16]. In this line, while changes in the diversity of gut microbiome are getting deeply studied, there is scarce data about diversity changes in blood microbiome among patients with viral hepatitis. In a recent study, a trend towards lower bacterial diversity with worse disease stage was described in the context of hepatitis B and D infection [18], which is consistent with what we observed in patients with HCV-related cirrhosis.

Regarding the beta diversity, which represents the number of overlapping taxa between samples, microbiome signature differences between compensated and decompensated patients were consistently found for phylum, class, and order taxonomic ranks, and corroborated by several indexes, with PCoA helping us to visualize these differences. In previous studies, separate clustering between patients with liver disease induced by viral hepatitis C and healthy controls was described in the gut [19]. However, similar to alpha diversity, information in the literature is sparse, and to our knowledge, this is the first report studying differences in beta diversity between different cirrhosis stages in blood.

We also detected an association between a higher relative abundance of Proteobacteria, Alphaproteobacteria, and Sphingomonadales and hepatic decompensation. Proteobacteria has been described as a potential microbial signature of dysbiosis and disease risk [20]. A study by Sun et al. documented that the phylum Proteobacteria, among others, was increased in patients with chronic B cirrhosis [17]. Alphaproteobacteria, a class rank belonging to the phylum Proteobacteria, is classified as one of the most diverse bacterial subdivisions influencing host-cell proliferation [21]. The order Sphingomonadales, which is part of the class Alphaproteobacteria, appears to be relatively more abundant in patients with hepatic decompensation. Bacteria from the Sphingomonadales order are known to have lipoxygenases that may have a vital role in bacteria-host signaling [22]. All these findings in the literature support the significant association with hepatic decompensation found in our study. In regards to the *Sphingomonadaceae* family, we found a trend of association between the relative abundance of this family and hepatic decompensation.

Dysbiosis in environments such as the gut or oral cavity often goes hand in hand with metabolic changes and disease progression [23]. Regarding the blood, we observed that metabolic changes also accompanied microbiome dysbiosis. The phylum Proteobacteria was directly associated with two metabolites in this study, ethanolamine and oleic acid. Firstly, ethanolamine, a source of nitrogen and carbon by diverse bacteria, has been associated with liver reparation processes in the damaged liver [24]. Secondly, oleic acid is a monounsaturated fatty acid with numerous beneficial properties in various animal and vegetable sources. In liver injuries, oleic acid recruitment from other tissues has been described as one of the rescue systems [25]. Some studies have observed its role in inhibiting chemotaxis and attenuating inflammation [26]. Additionally, we found an inverse correlation of the relative abundance of the phylum Proteobacteria, the order Sphingomonadales, and the family *Sphingomonadaceae* with the p-cresol, being found by different analytical platforms (GC-MS and LC-MS). P-cresol is originated by fermentation from the microbiota. Based on previous microbiological studies, since bacteria of the phylum Proteobacteria are Gram-negative, their cell envelope is more sensitive to p-cresol, so it is consistent that in the absence of this metabolite, these taxa could grow freely in the blood and promote dysbiosis [27]. Besides, Ikematsu *et al*., in forensic autopsy cases, suggested that low levels of p-cresol in blood could be explained by an accumulation of p-cresol in the liver in cases of liver diseases [28].

Additionally, it is crucial to note that this study was performed using a strict contamination-aware approach. Firstly, any potential contamination by needle with the skin microbiome was prevented by the high volume of blood withdrawn. Secondly, positive and negative controls were used throughout the sequencing pipeline in order to control any impact of the environment on the results. Thirdly, to avoid possible confusion with HIV coinfection, multivariate models including HIV status as a covariate were carried out, controlling the possible confounding effects of HIV-coinfection. Finally, adjustment for multiple comparison was performed, avoiding false significant associations. All of these approaches provide robustness to our data.

The following considerations should be taken into account for a correct interpretation of the results. Firstly, this study had a cross-sectional design; therefore, it does not allow us to determine the causal relationship of the findings. Secondly, the sample size was limited, which could restrict the statistical power to detect other significant differences in bacterial taxa between groups. Moreover, we could not carry out the statistical analysis separately for HCV-monoinfected and HIV/HCV-coinfected patients due to the limited sample size. Nevertheless, possible confusion with HIV coinfection was ruled out by including HIV status as a covariate in multivariate models.

## Conclusions

In conclusion, blood microbial diversity was significantly decreased in patients with decompensated HCV-related cirrhosis, who presented an enrichment of Proteobacteria, Alphaproteobacteria, and Sphingomonadales, compared to patients with compensated cirrhosis. The dysbiosis of the blood microbiome was accompanied by metabolomic changes. Further studies are needed to decipher the impact that blood microbial dysbiosis could have on these patients and strategies to correct it.

## Declarations

### Consent for publication

Not applicable

### Availability of data and materials

The datasets used and/or analyzed during the current study are available from the corresponding author upon reasonable request.

The raw sequences are publicly available at the European Nucleotide Archive repository (ENA; https://www.ebi.ac.uk/) under the accession number PRJEB65371.

## Data Availability

All data produced in the present study are available upon reasonable request to the authors.
The raw sequences are publicly available at the European Nucleotide Archive repository (ENA; https://www.ebi.ac.uk/) under the accession number PRJEB65371

## Acknowledgments

This study would not have been possible without the collaboration of all the patients, medical and nursery staff, and data managers who participated in the project. We want to particularly acknowledge the support of the HIV BioBank, integrated into the Spanish AIDS Research Network and all collaborating Centres, for the generous contribution with clinical samples for the present work (see **Appendix**). The HIV BioBank is supported by Instituto de Salud Carlos III, Spanish Health Ministry (Grant n° RD06/0006/0035, RD12/0017/0037 and RD16/0025/0019) as part of the Plan Nacional R + D + I and cofinanced by ISCIII-Subdirección General de Evaluación y el Fondo Europeo de Desarrollo Regional (FEDER)”. The RIS Cohort (CoRIS) is funded by the Instituto de Salud Carlos III through the Red Temática de Investigación Cooperativa en SIDA (RIS C03/173, RD12/0017/0018 and RD16/0002/0006) as part of the Plan Nacional R+D+I and cofinanced by ISCIII-Subdirección General de Evaluacion and the Fondo Europeo de Desarrollo Regional (FEDER).

## Authorship contribution

Funding body: MAJS, SR.

Study concept and design: MAJS.

Patients’ selection and clinical data acquisition: CD, VH, JB, JGG, LIS, ELH, AO, LPL.

Sample preparation, and biomarker analysis: OBK, AVB, AFR, DR, CB.

Statistical analysis and interpretation of data: OBK, MR, MAJS.

Writing of the manuscript: OBK, MAJS.

Critical revision of the manuscript for relevant intellectual content: JB, JGG, DR, CB, AFR, MR, SR.

Supervision and visualization: MAJS.

All authors read and approved the final manuscript.

## Ethical Approval statement

The study received the approval of the Research Ethics Committee of the Instituto de Salud Carlos III (CEI42_2020, CEI41_2014).

## Appendix The ESCORIAL study group

**Hospital General Universitario Gregorio Marañón (**Madrid, Spain): Cristina Díez, Luis Ibáñez, Leire Pérez-Latorre, Diego Rincón, Teresa Aldámiz-Echevarría, Vega Catalina, Pilar Miralles, Teresa Aldámiz-Echevarría, Francisco Tejerina, María C Gómez-Rico, Esther Alonso, José M Bellón, Rafael Bañares, and Juan Berenguer.

**Hospital Universitario La Paz/IdiPAZ (**Madrid, Spain): José Arribas, José I Bernardino, Carmen Busca, Javier García-Samaniego, Víctor Hontañón, Luz Martín-Carbonero, Rafael Micán, María L Montes-Ramírez, Victoria Moreno, Antonio Olveira, Ignacio Pérez-Valero, Eulalia valencia, and Juan González-García.

**Hospital Universitario Puerta de Hierro** (Madrid, Spain): Elba Llop and José Luis Calleja.

**Hospital Universitario Ramón y Cajal** (Madrid, Spain): Javier Martínez and Agustín Albillos.

**Fundación SEIMC/GeSIDA** (Madrid, Spain): Marta de Miguel, María Yllescas, and Herminia Esteban.

## Abbreviations

aAMR: Adjusted arithmetic mean ratio
AMR: Arithmetic mean ratio
ART: Antiretroviral therapy
CTP: Child-Turcotte-Pugh
ESI: Electrospray ionization
FC: Fold Change
FDR: False discovery ratio
GC-MS: Gas chromatography-mass spectrometry
GLM: Generalized linear model
HCC: Hepatocellular carcinoma
HCV: Hepatitis C virus
HIV: Human immunodeficiency virus
HVPG: Hepatic venous pressure gradient
INR: International normalized ratio
LC-MS: Liquid chromatography-mass spectrometry
MANOVA: Multivariate analysis of variance
OTU: Operative taxonomic Unit
PCoA: Principal coordinates analysis
PCR: Polymerase chain reaction

