## Supplementary material for "Altered blood microbiome in patients with HCV-related decompensated cirrhosis": Suplementary_file_1

### Supplementary data

### Supplementary File 1: Additional description of the methods section:

**Non-targeted metabolomics**

***Reagents and standards for metabolomics***

They have been used: acetonitrile (LC-MS grade, Sigma-Aldrich, Steinheim, Germany), formic acid (FA) (MS grade, Sigma-Aldrich, Steinheim, Germany), MilliQ® water (Millipore, Billerica, MA, USA), heptane (Sigma-Aldrich, Steinheim, Germany), methylterbutyleter (MTBE) (Sigma-Aldrich, Steinheim, Germany), pyridine (Sigma-Aldrich, Steinheim, Germany), O-methoxyamine hydrochloride (Sigma-Aldrich, Steinheim, Germany) and N,O-bis(trimethylsilyl) trifluoroacetamide (BSTFA) plus 1 % trimethylchlorosilane (TMCS) (Pierce Chemical Co, Rockford, IL, USA). Stearic acid methyl ester (C18:0 methyl ester) (Sigma-Aldrich, Steinheim, Germany) was used as internal standard for GC-MS. For reference masses purine, hexakis(1H,1H,3H-tetrafluoropropoxy)phosphazine (HP) and ammonium trifluoroacetate (TFA(NH_4_)) from Agilent (API-TOF reference mass solution kit) were used in LC-MS. In GC-MS, a FAME mix (fatty acid methyl esters, e.g. caprylic acid, capric acid, lauric acid, tridecanoic acid, myristic acid, myristoleic acid, pentadecanoic acid, palmitic acid, palmitoleic acid, heptadecanoid acid, stearic acid, elaidic acid, oleic acid, linoleic acid, arachidic acid, cis-11-eicosenoic acid, linolenic acid, behenic acid and erucic acid) was purchased from Supelco (Bellefonte, PA, USA).

***Sample preparation for metabolomics analysis***

On the day of the analysis, for GC-MS 100 µL of the corresponding methanolic aliquots were evaporated to dryness using a Speedvac Concentrator, followed by the addition of 10 μL of O-methoxyamine hydrochloride (15 mg/mL) in pyridine for methoximation. After gently vortexing, the vials were incubated in darkness at room temperature for 16 hours. Then, 10 μL of BSTFA with 1 % TMCS (v/v) were added, and samples were vortexed for 5 min. Silylation was carried out for one hour at 70°C, and finally 100 μL of C18:0 methyl ester (10 mg/L in heptane) were added as an internal standard, and samples were remixed by gently vortex. Six blank samples were prepared by the same procedure of extraction and derivatization. For LC-MS, 500 µL of MTBE were added to the samples to enhance the extraction of the lipophilic compounds. After gently vortexing (TissueLyser LT, 50 Hz, 10 min) and centrifugation (16000 g, 20 min, 4°C), the resulting supernatants were filtered through 0.22 µm nylon syringe filters and transferred into an analytical vial for their analysis.

Quality controls (QCs) samples are required at the beginning of the sequence to stabilize the system and throughout the analytical runs at periodic intervals to monitor signal variations across the time. For these reasons, individual QC samples were prepared independently for each analytical platform by pooling and mixing equal volumes of each corresponding sample. After gently vortex, the mixes were transferred to analytical vials.

***GC-EI-Q-MS fingerprinting*** (for using FiehnLib [1] and NIST 14 libraries)

GC system (Agilent Technologies 7890A) consisted in an autosampler (Agilent Technologies 7693) and an inert mass selective detector (MSD) with Quadrupole (Agilent Technologies 5975). Two μL of the derivatized sample were injected through a GC-Column DB5-MS (30 m length, 0.25 mm internal diameter, 0.25 μm film 95% dimethylpolysiloxane / 5% diphenylpolysiloxane) with a pre-column (10 m J&W integrated with Agilent 122-5532G). The flow rate of the helium carrier gas was set at 1 mL/min, and the injector temperature 250°C. The split ratio was 1:10 flow into a Restek 20782 deactivated glass-wool split liner. The temperature gradient was programmed at 60°C (held for 1 min), with a ramping increase rate of 10 °C/min up to 325°C. Finally, it was cooled down for 10 min before the next injection. The total analysis time was 37.5 min. The detector transfer line, filament source, and quadrupole temperature were respectively set at 290°C, 230°C, and 150°C. The electron ionization (EI) source was placed at 70 eV. The mass spectrometer operated in scan mode over a mass range of *m/z* 50–600 at a rate of 2 spectra per second. The method was retention time locked at 19.663 minutes (elution time of the internal standard). The analytical run was set up starting with the injection of C18:0 methyl ester (10 mg/L in heptane) and FAME mix (0.1 mg/mL in CH_2_Cl_2_) followed by four blanks, five QCs and then samples were analysed in a randomised order, where other QCs were injected between blocks of ten samples until the end of the run that terminated with the injection of the four blanks.

***LC-ESI-QTOF-MS fingerprinting***

The metabolic profile was achieved using a liquid chromatography system consisting of a degasser, a binary pump, and an autosampler (1290 infinity II, Agilent). Samples (0.5 µL) were applied to a reversed-phase column (Zorbax Extend C18 50 x 2.1 mm, 1.8 µm; Agilent), which was maintained at 60°C during the analysis. The system was operated at a flow rate of 0.6 mL/min with solvent A (H_2_O containing 0.1% FA) and solvent B (acetonitrile containing 0.1% FA). The gradient was 5% B (0–1 min), 5 to 80% B (1–7 min), 80 to 100% B (7–11.5 min), and 100 to 5% B (11.5–12 min). The system was finally held at 5% B for 3 min to re-equilibrate the system (15 min of total analysis time). Data were collected in positive and negative electrospray ionization (ESI) modes in separate runs using QTOF (Agilent 6550 iFunnel). The analyses were performed in both positive and negative ion modes in full-scan from *m/z* 50 to 1000. The capillary voltage was 3000 V and the nozzle voltage was 1000 V with a scan rate of 1.0 spectrum per second. The gas temperature was 250°C, the drying gas flow was 12 L/min, the nebulizer was 52 psi, the sheath gas temperature 370°C and the sheath gas flow 11 L/min. For positive mode, the MS-TOF parameters were as follows: fragmentor 175 V and octopole radio frequency voltage 750 V. For negative mode, the MS-TOF parameters included the following: fragmentor 250 V and octopole radio frequency voltage 750 V. During the analyses, two reference masses were used: 121.0509 (purine, detected *m/z* [C_5_H_4_N_4_+H]^+^) and 922.0098 (HP, detected *m/z* [C_18_H_18_O_6_N_3_P_3_F_24_+H]^+^) in positive mode and 112.9855 (TFA(NH_4_), detected *m/z* [C_2_O_2_F_3_(NH_4_)-H]^-^) and 966.0007 (HP+FA, detected *m/z* [C_18_H_18_O_6_N_3_P_3_F_24_+FA-H]^-^) in negative mode. The references were continuously infused into the system, enabling constant mass correction. Samples were analyzed in randomized runs, during which they were incubated in an autosampler at 4°C. The analytical runs for both polarities were set up starting with the analysis of ten QCs followed by the samples; a QC sample was injected between blocks of ten samples until the end of the run.

***Quality assurance***

After data reprocessing, the metabolic features were subsequently filtered. For GC-MS, 82 metabolites were detected. After filtering for a relative standard deviation (RSD) <50 and presence in at least 60% of the samples in each experimental group, 81 metabolites were selected for statistical analysis. For LC-MS, a normalization over the analysis time (injection order) was carried [2]. The detected features were 347 and 335 for ESI+ and ESI-, respectively. Out of them, 268 and 234 features fulfilled a value of RSD <50 and had presence in more than 60% of samples in each group.

***Metabolite identification***

The significant metabolites were identified. In GC-MS the identification was done based on FiehnLib [1] and NIST 14 libraries. In LC-MS the list of accurate masses was searched using the CEU Mass Mediator search tool (http://ceumass.eps.uspceu.es/; error ± 5 ppm) to obtain tentative identifications. Each of them was manually curated based on their MS adducts [3]. In the cases that it was applicable, the elution order was also considered to discard spurious identifications. Eventually, the biological role of each compound was evaluated, and unrelated identifications such as pesticides, drugs, or not possible chemical structures were excluded. The metabolites are reported in agreement with the criteria of the Metabolomics Standards Initiative [4, 5] with a confidence level grade 2 (putatively annotated compounds), which certitude is increased after manual curation of the final list.

**Supplementary File 2.** Association of richness and alpha diversity with cirrhosis decompensation.

| Index | **Taxonomic rank** | **AMR (95%CI)** | **p-value** | **aAMR (95%CI)** | **p-value** |
| --- | --- | --- | --- | --- | --- |
| Chao1 estimator | Phylum | 0.84 (0.74-0.96) | **0.012** | 0.85 (0.74-0.97) | **0.021** |
|  | Class | 0.85 (0.75-0.96) | **0.013** | 0.85 (0.74-0.97) | **0.019** |
|  | Order | 0.88 (0.76-1.04) | 0.121 | 0.90 (0.77-1.06) | 0.197 |
|  | Family | 1.16 (0.92-1.48) | 0.215 | 1.19 (0.94-1.54) | 0.,160 |
|  | Genus | 1.11 (0.85-1.50) | 0.461 | 1.21 (0.89-1.65) | 0.219 |
|  | Species | 0.96 (0.73-1.27) | 0.748 | 1.05 (0.79-1.41) | 0.753 |
| Shannon index | Phylum | 0.80 (0.69-0.92) | **0.003** | 0.80 (0.69-0.93) | **0.005** |
|  | Class | 0.91 (0.80-1.03) | 0.118 | 0.89 (0.78-1.01) | 0.066 |
|  | Order | 0.98 (0.91-1.07) | 0.667 | 0.97 (0.89-1.06) | 0.471 |
|  | Family | 0.97 (0.89-1.05) | 0.429 | 0.95 (0.87-1.04) | 0.275 |
|  | Genus | 0.96 (0.88-1.04) | 0.330 | 0.95 (0.87-1.03) | 0.196 |
|  | Species | 0.95 (0.88-1.04) | 0.273 | 0.94 (0.87-1.03) | 0.167 |
| Simpson index | Phylum | 0.81 (0.71-0.92) | **0.002** | 0.83 (0.72-0.95) | **0.006** |
|  | Class | 0.94 (0.80-1.03) | 0.186 | 0.93 (0.85-1.03) | 0.158 |
|  | Order | 1.00 (0.96-1.04) | 0.896 | 1.00 (0.96-1.04) | 0.898 |
|  | Family | 0.99 (0.96-1.03) | 0.586 | 0.99 (0.96-1.03) | 0.642 |
|  | Genus | 0.99 (0.96-1.02) | 0.405 | 0.99 (0.96-1.02) | 0.459 |
|  | Species | 0.99 (0.96-1.02) | 0.358 | 0.99 (0.96-1.02) | 0.419 |

**Statistics:** P-values were calculated by generalized linear models under a gamma distribution unadjusted (AMR) and adjusted by HIV coinfection. Results are shown as arithmetic mean ratio and 95% confidence interval). Cirrhosis decompensation: CTP≥7 and CTP<7.

**Abbreviations**: AMR, arithmetic mean ratio; aAMR, adjusted arithmetic mean ratio; CI, confidence interval.

**Supplementary File 3.** Association of beta diversity with cirrhosis decompensation

|  | **Weighted Unifrac**  **p-value** | **Adjusted Weighted Unifrac**  **p-value** | **Bray-Curtis**  **p-value** | **Adjusted Bray-Curtis**  **p-value** | **Jaccard**  **p-value** | **Adjusted Jaccard**  **p-value** |
| --- | --- | --- | --- | --- | --- | --- |
| **Phylum** | 0.078 | 0.099 | 0.100 | 0.102 | **0.049** | **0.046** |
| **Class** | **0.040** | **0.033** | **0.017** | **0.018** | **0.028** | **0.023** |
| **Order** | **0.016** | **0.019** | **0.048** | **0.048** | 0.072 | 0.071 |
| **Family** | 0.057 | 0.064 | 0.175 | 0.173 | 0.252 | 0.246 |
| **Genus** | 0.158 | 0.230 | 0.438 | 0.429 | 0.568 | 0.566 |
| **Species** | 0.188 | 0.162 | 0.244 | 0.242 | 0.338 | 0.336 |

**Statistics:** P-values were calculated by permutational multivariate analysis of variance using distance matrices. Adjusted models were adjusted by HIV coinfection.

**Supplementary File 4.** Boxplots representing the relative abundance of bacterial taxa significantly associated with hepatic decompensation. **Abbreviations:** CTP, Child-Turcotte-Pugh.

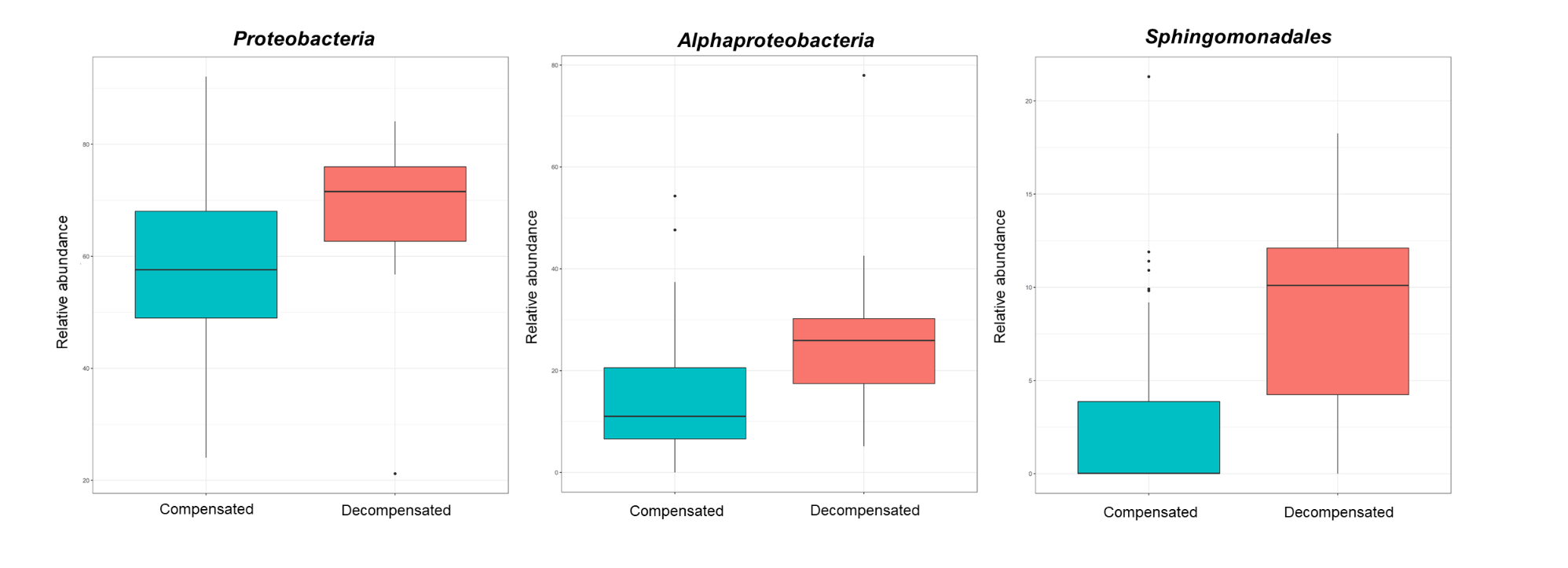

**Supplementary File 5.** Summary of significant correlations between significant bacterial taxa and metabolomics data using Spearman correlation analysis.

|  | | *Proteobacteria* | | | *Alphaproteobacteria* | | | *Sphingomonadales* | | |
| --- | --- | --- | --- | --- | --- | --- | --- | --- | --- | --- |
|  |  | r | p | q | r | p | q | r | p | q |
| GC-MS | Ethanolamine | 0.330 | **0.005** | **0.115** | 0.145 | 0.226 | 0.886 | 0.102 | 0.399 | 0.674 |
|  | Oleic acid | 0.302 | **0.004** | **0.115** | 0.071 | 0.511 | 0.886 | 0.248 | **0.020** | 0.260 |
|  | p-Cresol | -0.308 | **0.006** | **0.115** | -0.230 | 0.042 | 0.740 | -0.354 | **0.001** | **0.112** |
| LC-MS  ESI+ | 5S-HETE di-endoperoxide | 0.163 | 0.134 | 0.757 | 0.060 | 0.582 | 0.954 | 0.328 | **0.002** | 0.267 |
|  | PC(16:0/9:0(CHO)) | 0.322 | **0.003** | 0.678 | 0.118 | 0.277 | 0.954 | 0.052 | 0.635 | 0.905 |
|  | LPE (20:4) | 0.012 | 0.913 | 0.966 | -0.063 | 0.564 | 0.954 | -0.307 | **0.004** | 0.267 |
| LC-MS  ESI- | 3-OH-isovaleric acid | 0.310 | **0.004** | 0.214 | 0.153 | 0.163 | 0.993 | -0.013 | 0.908 | 0.977 |
|  | 3-OH-butyric acid | 0.304 | **0.004** | 0.214 | 0.150 | 0.169 | 0.993 | 0.098 | 0.368 | 0.977 |
|  | p.Cresol | -0.150 | 0.170 | 0.999 | -0.259 | **0.017** | 0.920 | -0.367 | **0.001** | **0.122** |

**Statistics:** Correlations were calculated by non-parametric Spearman correlation analysis. Q-values were calculated by false discovery rate using Benjamini and Hochberg correction,

**Abbreviations**: GC-MS, gas chromatography mass spectrometry; LC-MS, liquid chromatography mass spectrometry; p, p-value; q, q-value; ESI, electrospray ionization.
